# Cellular and Transcriptional Signature of the Nasal Mucosa is Associated with Susceptibility to Pneumococcal Carriage in Older Adults

**DOI:** 10.1101/2023.11.16.23298619

**Authors:** Britta C. Urban, André N. A. Gonçalves, Dessi Loukov, Fernando M. Passos, Jesús Reiné, Patrícia Gonzalez-Dias, Carla Solórzano-Gonzalez, Elena Mitsi, Elissavet Nikolaou, Daniel O’Connor, Andrea M. Collins, Hugh Adler, Jamie Rylance, Stephen B. Gordon, Simon P. Jochems, Helder I. Nakaya, Daniela M. Ferreira

**Affiliations:** Oxford Vaccine Group, University of Oxford, Oxford, United Kingdom; Clinical Sciences, Liverpool School of Tropical Medicine, Liverpool, United Kingdom; NIHR Oxford Biomedical Research Centre, Oxford, UK; Infection, Immunity and Global Health, Murdoch Children’s Research Institute, Parkville, Victoria, Australia; Department of Microbiology and Immunology at the Peter Doherty Institute for Infection and Immunity, The University of Melbourne, Parkville, Victoria, Australia; Malawi-Liverpool-Wellcome Clinical Research Programme, Blantyre, Malawi; Department of Parasitology, Leiden University Centre for Infectious Diseases, Leiden University Medical Centre, Leiden, the Netherlands; Hospital Israelita Albert Einstein, São Paulo, Brazil; Department of Clinical and Toxicological Analyses, School of Pharmaceutical Sciences, University of São Paulo, São Paulo, Brazil

**Author notes:** These authors contributed equally (Britta C. Urban, André N. A. Gonçalves). Corresponding authors: Britta Urban, Daniela Ferreira. Contributions: Obtained funding: SBG, DMF Study design: HA, DMF, JRy Obtained ethical approval: HA, JRy Data collection: HA, AC, JR, SJ, CS, EM, EN Data analyses: DL, FMP, BU, ANAG, PGD Manuscript preparation: BU, ANAG, DO, DMF, HN, SJ. All authors contributed to the manuscript and approved the final version submitted for publication.

## Abstract

*Streptococcus pneumoniae* colonization in the upper respiratory tract is linked to pneumococcal disease development, predominantly affecting the very young and older adults. As the global population ages and comorbidities increase, there is a heightened concern about this infection. We investigated the immunological responses of older adults to pneumococcal controlled human infection by analysing the cellular composition and gene expression in the nasal mucosa. Our comparative analysis with younger adults revealed distinct gene expression patterns in older individuals susceptible to colonization, highlighted by neutrophil activation and elevated levels of CXCL9 and CXCL10. Unlike younger adults challenged with pneumococcus, older adults did not show recruitment of monocytes into the nasal mucosa following nasal colonization. These findings suggest age-associated cellular changes, in particular enhanced mucosal inflammation, that may predispose older adults to pneumococcal colonization. If similar changes are observed in the lung of susceptible older adults, these may explain the increased risk of pneumococcal disease in vulnerable populations.

## Introduction

*Streptococcus pneumoniae* is the primary cause of bacterial pneumonia and can reside in the upper respiratory tract without causing symptoms. Colonization with pneumococcus is a prerequisite of pneumococcal disease, with higher colonization levels increasing the risk of bacteria entering the lung^1^. Invasive pneumococcal disease and community-acquired pneumonia are a leading cause of death worldwide. Community-acquired pneumonia affects young children (below five years) and older adults disproportionally^2–6^. With an aging population and rising prevalence of comorbidities such as COPD, obesity, diabetes, and asthma, the incidence of pneumonia is predicted to grow.

Our pneumococcal controlled human infection model is an ideal model to investigate molecular and cellular mechanisms that contribute to protection or susceptibility to colonization by pneumococci^7^. Using this model in young adults, we have shown that both innate and adaptive immune responses at the mucosa were associated with protection from colonization. We have also shown that colonization is typically established within a day after nasal exposure, ultimately prompting a local immune response that clears the pneumococci^8^. This response included the activation of neutrophils and a likely CCL2-mediated influx of monocytes, resulting in bacterial clearance through processes like phagocytosis and the release of reactive oxygen species^9^. Younger individuals resistant to colonization have more mucosal-associated invariant T cells (MAIT) in their nasal mucosa and in the peripheral circulation exhibit elevated TNFα and IFNγ levels when exposed to Spn6B^10^. In addition, young study participants protected from colonization showed a higher frequency of pre-existent peripheral blood memory B cells specific to the polysaccharide of the inoculated strain^11^. Moreover, colonization itself is an immunising event that results in increased antigen-specific B cells in circulation and higher levels of serum antibodies^10,11^. Although the impact of the SARS-CoV-2 pandemic increased the number of studies investigating mucosal immune, they remain rare in older adults. As a consequence, whether similar mechanisms are involved in protection and susceptibility of older adults from pneumococcus infection is not known.

Older adults face a higher risk of severe pneumococcal disease than younger adults. While other factors such as co-morbidities, increased lung microaspiration and viral co-infections may contribute to this increased risk^12^, altered immune responses due to aging also play an important role^13–15^. In our study, when we exposed healthy older adults (over 50 years) to pneumococcus, their colonization rates and densities mirrored that of younger adults^16^. However, their immune response differed – those who became colonized did not exhibit the expected boost in specific IgG against the inoculated strain capsular polysaccharide (CPS), as seen in younger adults^16^. In addition, in those participants who were challenged and did not become colonized, IgG levels to the CPS decreased a month after challenge. Re-challenging them with homologous pneumococcus 6B up to a year later did not protect older adults from colonization, unlike in younger adults where re-challenge conferred protection^7,16^. Together, these findings suggest a diminished immunizing effect of colonization in older adults, potentially increasing their vulnerability to severe pneumococcal disease even with lower colonization rates^17^.

To gain a better understanding of mechanisms underlying the lack of protection, we investigated here the changes in cellular composition, inflammatory status and gene expression patterns of nasal cells from older adults before and during experimental human pneumococcal colonization. We then compared these changes with those from younger adults^9,18^. Older but not younger adults susceptible to colonization displayed a distinct pattern of activated gene pathways associated with neutrophil degranulation and increased concentrations of CXCL9 and CXCL10 in the nasal mucosa at baseline. Our results open the possibility of predicting susceptibility to pneumococcal colonization and greater risk of pneumococcal disease in older adults which would facilitate better prevention and treatment.

## Results

### Distinct gene expression patterns in the nasal mucosa of older adults linked to carriage susceptibility

We previously conducted an experimental human pneumococcal challenge trial in healthy, older adults^16^ during which we collected nasal lining fluid and nasal microbiopsies to determine soluble immune mediators, nasal cell phenotype and transcriptome from a subset of study participants. An overview of the sample schedule relative to inoculation with serotype 6B (Spn6B) for older adults is provided in Supplementary Fig. 1. No demographic differences were seen between study participants susceptible (n=17) or protected (n=24) from colonization (Supplementary Table 1).

We first investigated which immune response pathways were enriched in protected and susceptible older adults before (day –5) and after (day 2 and day 9) pneumococcal challenge using gene set variation analysis (GSVA) (Fig. 1a, Supplementary Table 2). Based on Euclidean distance clustering, gene sets fell into three groups with different enrichment patterns over time and carriage status. Group I gene sets were associated with innate immune responses and cytokine signalling including pathways associated with “Antimicrobial peptides”, “Neutrophil degranulation”, “DAP12 interactions” and “Toll-like receptor cascades”. They remained higher across all time points in susceptible older adults but were not induced in protected older adults. By contrast, group II gene sets were largely associated with antibody-mediated processes such as “Complement Cascade” and “Fc gamma receptor dependent phagocytosis”. These pathways were predominantly enriched at baseline (day – 5) in protected older adults and induced in susceptible older adults by day 9. Group III gene sets associated with antigen presentation and T cell activation and were induced in susceptible study participants on day 9.

**Fig. 1.**
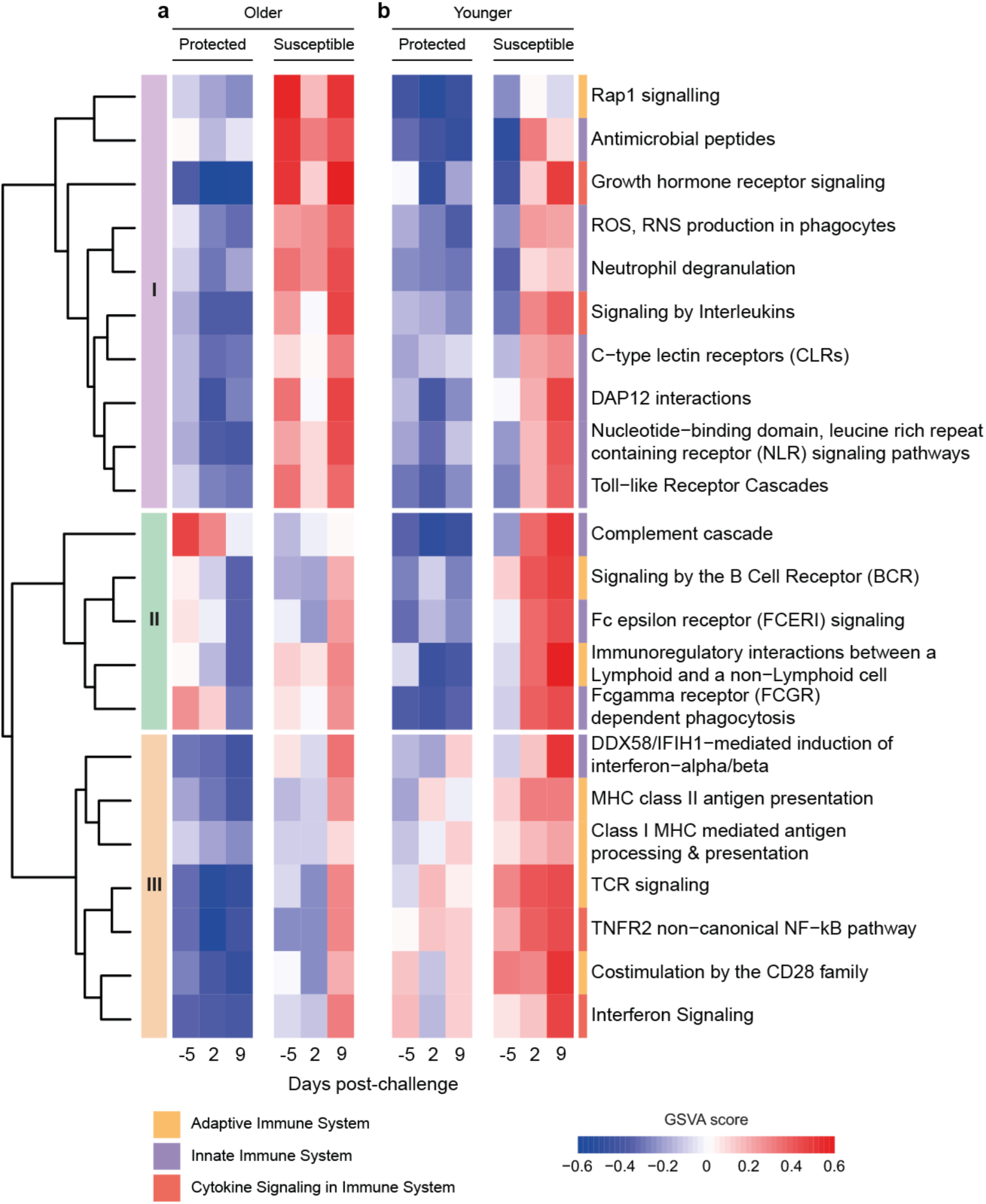
Gene expression analysis of nasal cells showed distinct patterns in older adults susceptible to carriage before pneumococcal challenge. **a**, Gene set variance analysis (GSVA) of immune response pathways of nasal cells collected before (day –5) and after (day 2, day 9) pneumococcal challenge of older and b, younger study participants who were protected (carriage-negative) or susceptible (carriage-positive). Based on Euclidean distance calculation, we identified three groups: group I contains gene sets associated predominantly with innate immune responses, group II and III are enriched for gene

We then asked whether similar gene pathway patterns were evident in younger adults before pneumococcal challenge using a control group from our previously published studies (protected n=6, susceptible n=8, age < 50 years)^9,18^. In younger adults, GSVA of the nasal cell transcriptome showed no difference in the regulation of group I gene or group II gene sets at baseline, between protected and susceptible study participants. However, gene pathways in both groups were rapidly induced after pneumococcal challenge in susceptible younger adults indicating activation of innate and antibody-mediated immune responses (Fig 1b, Supplementary Table 3). By contrast, activation of gene sets in group III associated with “TCR signalling”, “Costimulation by the CD28 family” and MHC class I and class II antigen processing were enriched in susceptible younger adults at baseline. In summary, gene expression patterns in the nasal mucosa were fundamentally different between susceptible younger and older adults, with susceptible older adults exhibiting a unique gene set signature associated with an activated innate immune response profile before pneumococcal challenge.

### Co-expression analysis of genes reveals upregulation of neutrophil degranulation pathways in the nasal mucosa of susceptible older adults

Co-expression analysis allows identification of genes which are co-regulated during cellular processes or different functional states and which are shared within a sample group. We therefore investigated which genes were co-expressed in susceptible and protected older adults using CEMiTool^19^ across all three timepoints. Co-expression analysis identified three modules (M1-M3, Fig. 2a) with module M1 genes co-expressed in protected and M2 genes co-expressed in susceptible older adults at baseline. Notably, genes within the M2 module were enriched for the Reactome pathways “Neutrophil Degranulation”, “Immunoregulatory interactions between a Lymphoid and a Non-lymphoid Cell” and “Signalling by Interleukins” (Supplementary Fig. 2) which were also among upregulated gene sets in susceptible older adults identified by GSVA.

**Fig. 2.**
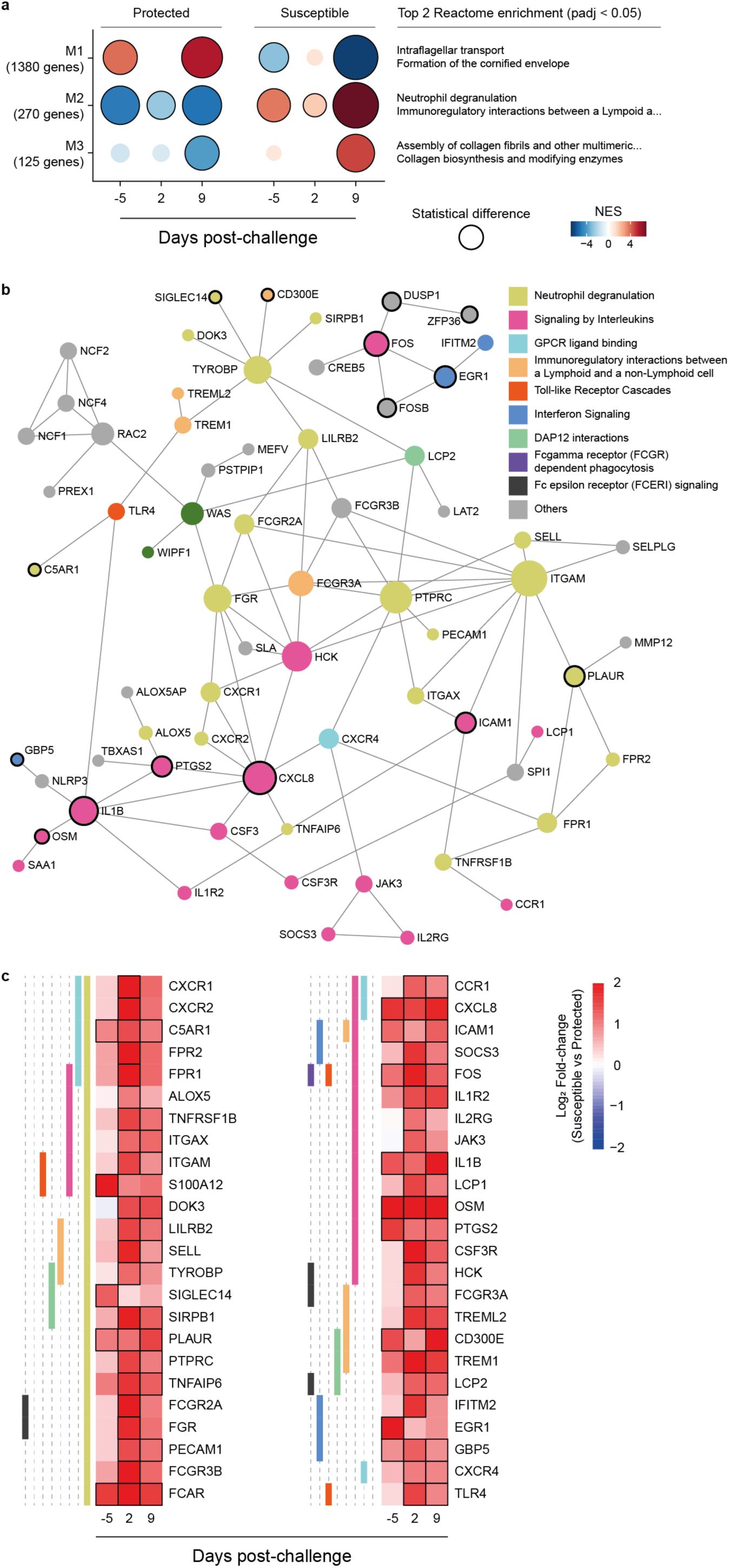
Co-expression analysis of nasal cell transcripts using protection-status and timepoints samples for study participants. **a**, Co-expressed genes were determined by CEMiTool (Co-Expression Module identification Tool), assessing normalised gene expression profile using samples of protection-status (susceptible and protected), and across the timepoints (baseline (day –5), day 2, and day 9 post-challenge) and Over-Representation Analysis (enrichr function from R package clusterProfiler) to determine the biological function of each module genes using Reactome database (level 3 gene sets). Gene Set Enrichment Analysis (GSEA) tool and z-score values for each gene across all timepoints were used to determine whether module genes were positively (positive NES values) or negatively (negative NES values) enriched. **b,** Protein-protein interaction network using 216 M2 modules and differentially expressed genes (susceptible vs protected samples) at at least one timepoint. Genes are colour coded by Reactome pathway and those which were overexpressed at baseline are indicated by a black border. StringDB was used to determine protein-protein interactions using default parameters, a confidence score cut-off of 900 and not required experimental evidence (NetworkAnalysit.ca webtool). **c,** Heatmap showing log2 fold-change of overexpressed genes within each pathway before and after pneumococcal challenge (susceptible vs protected samples). Criteria of differentially expressed genes were absolute log2 fold-change > 1 and non-adjusted p value < 0.05.

We then identified genes in the M2 module linked to carriage susceptibility by examining which ones were differentially expressed between susceptible and protected older adults. Module M2 identified 270 co-expressed genes (Supplementary Fig. 2) of which 216 were differentially expressed at at least one timepoint (Supplementary Table 4). Network analysis of genes overexpressed in susceptible study participants before challenge (Fig. 2b) revealed a more granular picture of interlinked immune response pathways. Genes overexpressed within the neutrophil degranulation pathway at baseline in susceptible study participants included *SIGLEC14* (2.42-fold, p=0.037), encoding a sialic acid binding protein and *PLAUR* (2.13-fold, p=0.012), encoding the Plasminogen Activator Urokinase receptor. Notably, the expression of PLAUR is upregulated by immune activation and the soluble form has been proposed as a marker for chronic inflammation^20^. Both, *SIGLEC14* and *TREM1* (4-fold, p=0.0028) which is induced two days after challenge signal through the DAP12 (*TYROBP*, 2.3-fold, p=0.027)) complex resulting in enhanced survival of immune cells and increased production of pro-inflammatory cytokines^21,22^. In a mouse model of pneumococcal pneumonia, TREM1 is critical for migration of neutrophils to the lung and enhanced early response of alveolar macrophages^23^. Of the Fc receptors, *FCAR* encoding the FcαR was 3.13-fold overexpressed at baseline (p=0.0078) whereas genes encoding Fcγ receptors were induced on days 2 and 9 after pneumococcal challenge (Fig. 2c). Of the M2 cytokine and chemokine genes, *IL1B* (2.6-fold, p=0.016) and *CXCL8* (3.4-fold, p=0.0031) showed higher expression in nasal cells of susceptible compared with protected older adults at baseline.

### Increased baseline expression of CXCL9 and CXCL10 in the nasal mucosa of susceptible older adults

Given the increased expression of genes associated with innate immune and cytokine responses in susceptible older adults, we asked whether transcriptional changes are at least in part reflected at the protein level in the nasal mucosa (Fig. 3 and Supplementary Fig. 3). The median concentration of the chemokine CXCL10 (IP10) was 2.27-fold higher before (day –5) and 3.4-fold higher on day 2 in nasal lining fluid after challenge in susceptible compared with protected study participants. Likewise, the concentration of the related chemokine CXCL9 was 1.16-fold increased at baseline (day –5) in susceptible older adults (Fig. 3a). Expression of the genes encoding these two chemokines were also increased in susceptible compared with protected older adults at baseline only (Fig. 3b, *CXCL9* 2.1-fold, p=0.021); *CXCL10* 2.6-fold, p=0.003). These data are reminiscent of younger adults with an asymptomatic viral infection or receiving Live Attenuated Influenza Vaccine (LAIV), who showed high baseline levels of CXCL10 and increased susceptibility to pneumococcal colonization upon challenge^9^. As with younger adults, there were no differences between CCL2 concentrations between susceptible and protected older adults. Unlike in younger adults, we did not observe an association between CCL2 concentration and the frequency of monocytes in the nasal mucosa (Supplementary Fig. 4). However, *CCL2* gene expression was 4 times higher in susceptible compared to protected older adults at day 9 after challenge suggesting that monocyte recruitment might be delayed. In addition, the concentration of IL-15, a cytokine which regulates T cell proliferation and activation, gradually increased in protected study participants after challenge and was 1.3-fold higher at day 9 compared to susceptible study participants. There were no statistically significant differences between protected and susceptible study participants before and after pneumococcal challenge for all other cytokines analysed (Supplementary Fig. 3a).

**Fig. 3.**
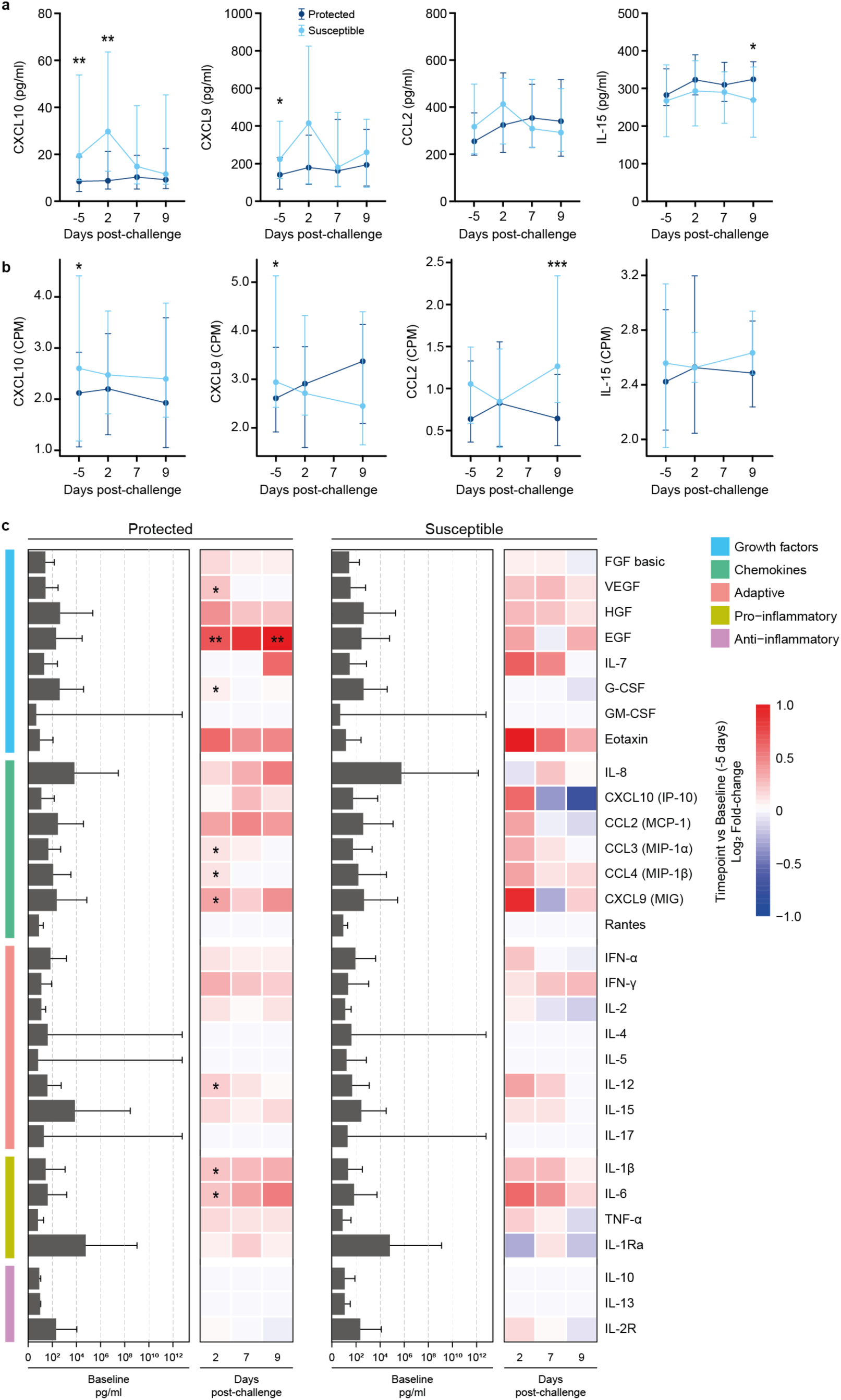
Changes in cytokine concentration in the nasal lining fluid of older adults experimentally challenged with *S. pneumoniae* 6B. **a**, Line graphs showing the median and interquartile range of cytokine concentrations in the nasal lining fluid in older study participants before (day –5) and after (day 2, day 7 and day 9) pneumococcal challenge in susceptible (light blue line, n=22) and protected (dark blue line, n=35) study participants. In protected study participants, the concentration of IL-15 was higher at day 9 (Mann-Whitney U test, * p=0.0344), and in susceptible study participants concentration of CXCL10 (IP-10) was higher at day –5 and day 2 (Mann-Whitney U test, * p=0.00116, ** p=0.0044) and the concentration of CXCL9 (MIG) was higher at day –5 (Mann-Whitney U test, * p=0.0419). No differences were observed for CCL2 (MCP-1). **b,** Line graphs showing the median and interquartile range of cytokine normalised gene expression (CPM= counts per million) in older study participants before (day –5) and after (day 2, and day 9) pneumococcal challenge in susceptible (light blue line, n=18) and protected (dark blue line, n=24) study participants. In susceptible study participants normalised gene expression of CXCL10 (IP-10) and CXCL9 (MIG) was higher at day –5 (DESeq2 Wald test, * p=0.00303, * p=0.0210, respectively), and CCL2 was higher at day 9 (DESeq2 Wald test, *** p=0.000218). No differences were observed for IL-15. **c,** Bar graph showing the level of cytokines at baseline and heatmap depicting log2 fold-change at day 2, 7, and 9 relatives to baseline (day –5) samples. The cytokines were grouped based on activity, adaptive, anti-inflammatory, chemokines, growth factors, and pro-inflammatory responses. The statistical test performed was the non-parametric Wilcoxon test. One asterisk (*) indicates p-value < 0.05, and two asterisk (**) for p-value < 0.01.

When considering induction of cytokines within each group of susceptible or protected study participants (log_2_ fold change for day 2, 7 and 9 against baseline), we observed responses to pneumococcal challenge in the nasal mucosa of protected but not susceptible study participants (Fig. 3c and Supplementary Table S5). EGF significantly increased from baseline in protected study participants on days 2 and 9 and VEGF and G-CSF increased on day 2. In protected study participants, the concentration of the monocyte chemoattractant MIP1α and MIP1β, as well as CXCL9 and IL12 increased from baseline to day 2. The pro-inflammatory cytokines IL1β and IL6 were also induced. This is in contrast to observations in protected younger adults^8,9^ who showed rapid induction of nasal cytokines which peaked within 24 hours after challenge but were undetectable by 48h (day 2) indicating that protected older adults may show a slight delay in nasal cytokine responses to pneumococcal challenge. Cytokine levels at baseline did not correlate with age in study participants (Supplementary Fig. 3b).

### Lack of monocyte recruitment in susceptible older adults after pneumococcal challenge

The nasal cell gene expression profiles suggested that older adults susceptible to pneumococcal colonization had higher levels of innate immune cell activation compared to protected older adults. Granulocytes are abundant in the nasal mucosa while monocytes are sparse with the later rapidly recruited after challenge in younger adults^9,24^.

We analysed the cellular composition of nasal cells by flow cytometry from nasal microbiopsies which were obtained in parallel to those used for transcriptome analysis. The overall number of leukocytes, granulocytes and monocytes as a ratio to epithelial cells^9,25^ were comparable between younger and older adults before pneumococcal challenge (Fig. 4a). After challenge, there were no differences in the proportion of leukocytes or granulocytes over the following 29 days between susceptible and protected study participants in either younger or older adults (Fig. 4b, c). In contrast to younger adults^9^, monocytes were not recruited into the nasal mucosa of susceptible older adults in response to pneumococcal challenge (Fig. 4d). These results are in line with the limited and variable induction of CCL2 (MCP1) in susceptible older adults (Fig. 3a).

**Fig. 4.**
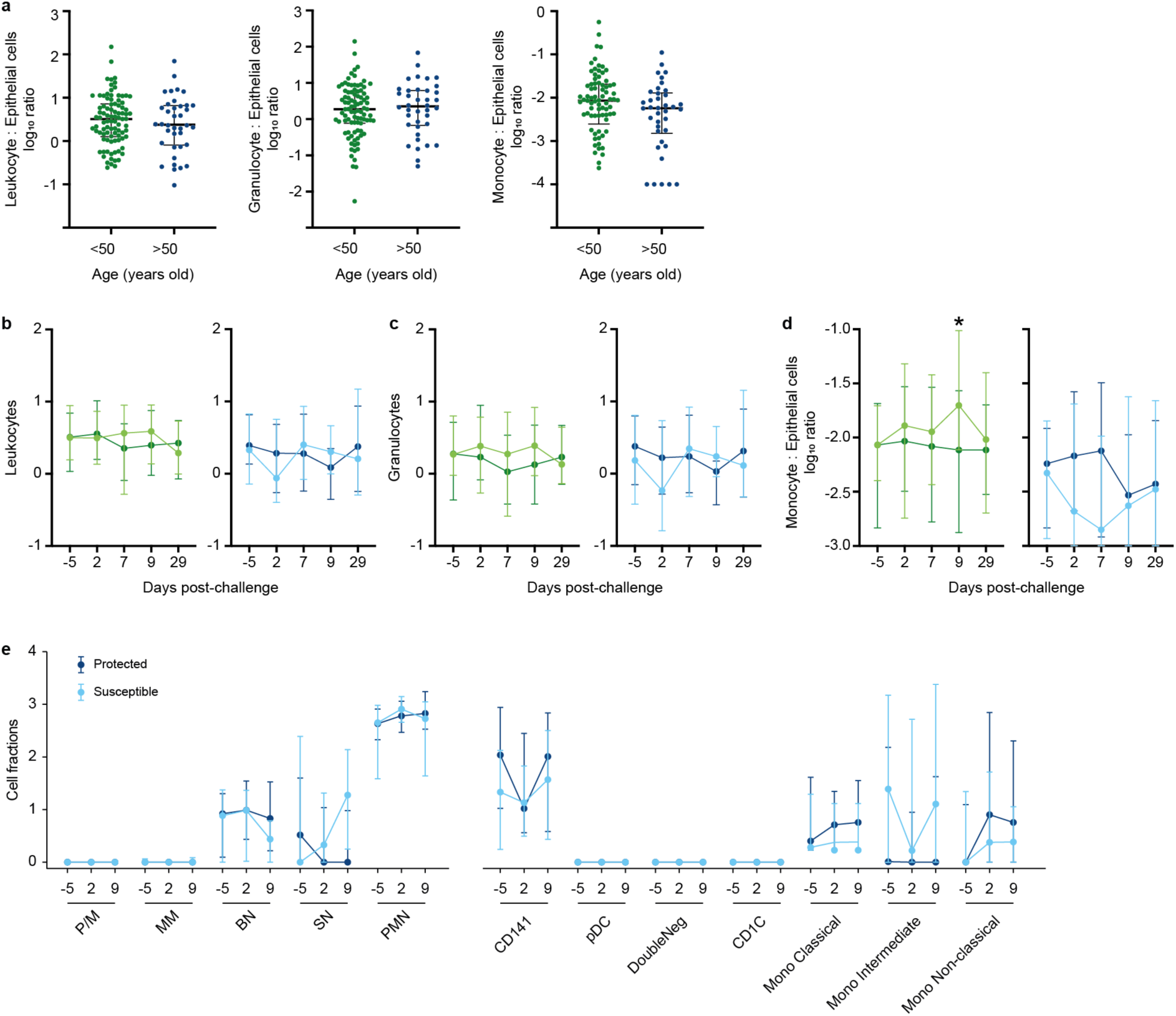
Recruitment of monocytes into the nasal mucosa of susceptible older adults is impaired. **a**, In nasal micro-biopsies, the number of leukocytes, granulocytes and monocytes was comparable between younger (green, n=91) and older adults (blue, n=38) before pneumococcal challenge (day –5). Cell numbers of each leukocyte subset were normalised to epithelial cells as a ratio to account for differences in the total number of cells obtained for each biopsy. **b-d,** Line graphs showing median and interquartile range of leukocytes, (b), granulocytes (c), and monocytes (d) in younger and older study participants before (day –5) and after (day 2, day 7, day 9 and day 29) pneumococcal challenge in susceptible (young: light green n= 28; old: light blue line, n=22) and protected (young: dark green, n=39; old: dark blue line, n=35) study participants. The proportion of monocytes increased at day 9 after challenge in susceptible compared to protected younger adults (Mann Whitney U test, * p=0.0122). This recruitment of monocytes into the nasal mucosa was not observed in susceptible older adults (n=15). **e**, Deconvolution analysis of granulocytes and myeloid cells using CIBERSORTx. Using normalised gene expression (log2 CPM (Counts Per Million)) and granulocytes and monocytes databases to determine the estimated cell population proportion. P/M (promyelocytes/myelocytes); MM (metamyelocytes); BN (band neutrophils); SN (segmented neutrophils) and PMN (polymorphonuclear). CD141 (CD141+ dendritic cells), pDC (plasmacytoid dendritic cells), double negative (CD1c-CD141-dendritic cells), CD1C (CD1c+ dendritic cells), mono (monocytes).

To gain a better understanding of granulocyte and monocyte subsets beyond the limited number of surface markers used during flow cytometry, we applied deconvolution analysis of mixed cell populations obtained in our bulk transcriptome data. Based on this analysis, mature neutrophils were the most abundant granulocyte population in the nasal mucosa independent of carriage status and timepoint (Fig. 4e, Supplementary Table S6). Myeloid cells were more heterogenous with CD141+ cDC1 the most abundant population in both susceptible and protected study participants. Intermediate monocytes appeared more abundant in susceptible participants at baseline. Intermediate monocytes display an activated phenotype and are generally associated with more inflammatory conditions. Notably, they were enriched in the nasal mucosa of older adults but not younger adults infected with Influenza A virus and correlated with age^24^.

### Activation of neutrophils is associated with susceptibility to carriage in older adults

Since susceptible older adults showed enrichment of gene pathways associated with neutrophil degranulation before challenge (Fig 1 and Fig. 2), we investigated the activation of granulocytes and monocytes before (day –5) and after (days 2, 7, 9, 29) pneumococcal challenge. The expression level of CD16 – but not CD66b or HLA DR – was consistently lower on granulocytes of older adults susceptible to pneumococcal colonization with significant lower levels observed at baseline and day 7 after challenge (Fig. 5a). Although FcγRIIIB can be stored in intracellular vesicle before rapid deployment onto the cell membrane in response to activation^26,27^, higher expression of CD16 on granulocytes positively correlated with gene expression levels for *FCGR3B* in protected but not susceptible older adults at baseline (Fig. 5b). CD16 is rapidly cleaved from the surface following activation of granulocytes indicating that these cells may have been already activated before challenge in older adults susceptible to pneumococcal carriage.

**Fig. 5.**
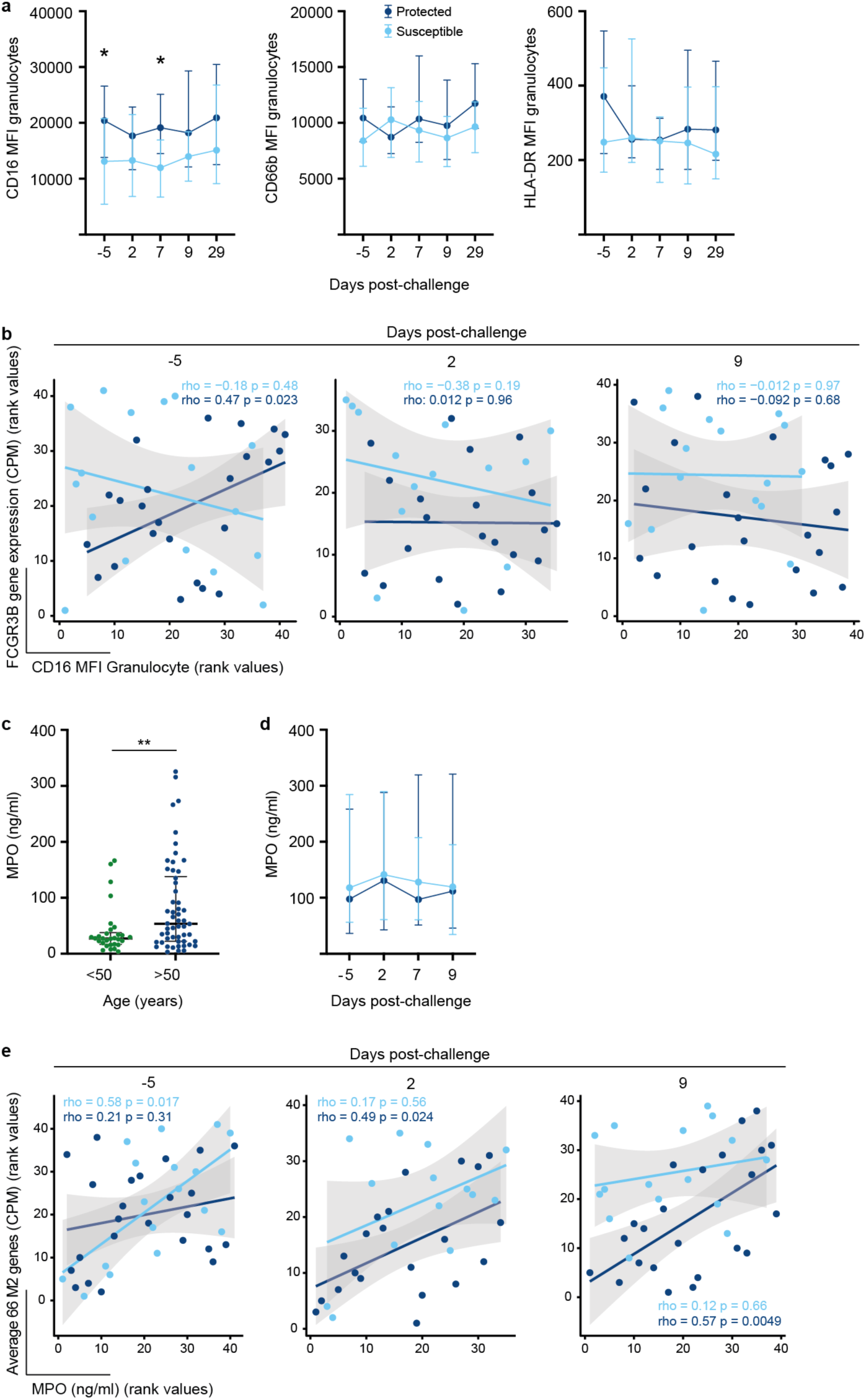
Granulocytes are activated in older adults susceptible to pneumococcal colonisation. **a**, Line graph (median and interquartile range) of expression levels of CD16, CD66b and HLA-DR on granulocytes before (day –5) and after challenge (day 2, day 7, day 9 and day 29) in protected (dark blue, n=34) and susceptible (light blue, n=22) older adults. The expression level of CD16 was significantly lower on nasal granulocytes at baseline and day 7 (Mann Whitney U test, baseline: p=0.034, day 7: p=0.014). **b**, Correlation between expression levels of FCGR3B gene and CD16 Granulocyte levels in nasal lining fluid at baseline (day –5), day 2 and day 9 in protected and susceptible study participants. **c**, At baseline (day –5), the concentration of MPO (Myeloperoxidase) in nasal lining fluid in older adults (n=54) was higher than in younger adults (n=30). Shown are dot plots with median and interquartile range, Mann Whitney U test ** p=0.0085. **d,** The concentration of MPO did not change before or after pneumococcal challenge in older study participants who became susceptible (light blue, n=22) or remained protected (dark blue, n=33). Line graph showing median and interquartile range of MPO concentration before and after inoculation with pneumococcus. **e**, Correlation between expression levels of 66 M2 module genes associated with the Reactome pathway “neutrophil degranulation” and MPO concentration in nasal lining fluid at baseline (day –5), day 2 and day 9 in protected and susceptible study participants.

Opsonisation, degranulation and radical oxygen species production are important defence mechanisms against colonization with pneumococcus^28^. To determine whether neutrophil degranulation increased after challenge with pneumococcus as we reported for younger adults^9^, we measured the concentration of myeloperoxidase (MPO), a soluble marker for degranulation of azurophil neutrophil granules, in nasal lining fluids. Before challenge, the concentration of MPO in the nasal lining fluid was 2-fold higher in older adults compared to younger adults (Fig. 5c). Unlike carriage-positive younger adults, levels of MPO in the nasal lining fluid did not increase after challenge in susceptible older adults (Fig 5d).

Next, we asked whether expression levels of 66 genes associated with the neutrophil degranulation pathway within the M2 module correlated with the MPO concentration in nasal lining fluid (Fig. 5e). MPO levels positively correlated with expression levels of neutrophil degranulation M2 module genes in susceptible older adults before but not after challenge suggesting that granulocytes were refractory to further activation. By contrast, in older adults protected from carriage, neutrophil degranulation genes within the M2 module correlated with MPO levels after but not before challenge implying that granulocytes became activated and contributed to clearance of pneumococci from the nose as we reported previously^8^. Together these data indicate that neutrophils in susceptible older adults are in a state of activation before challenge with limited responses to pneumococci after challenge.

### Activation of monocytes is associated with susceptibility to carriage in older adults

Given the lack of monocyte recruitment, we analysed expression levels of CD14, CD16 and HLA-DR on monocytes. Expression levels of CD14 but not HLA-DR or CD16 were significantly lower on monocytes before challenge (day –5) in susceptible compared to protected study participants (Fig. 6) indicating a dominance of intermediate monocytes in the nasal mucosa of susceptible older adults in agreement with deconvolution analysis of nasal transcriptome data (Fig. 4e). In protected older adults, the expression levels of CD16 increased on Day2 after challenge (Fig. 6).

**Fig. 6.**
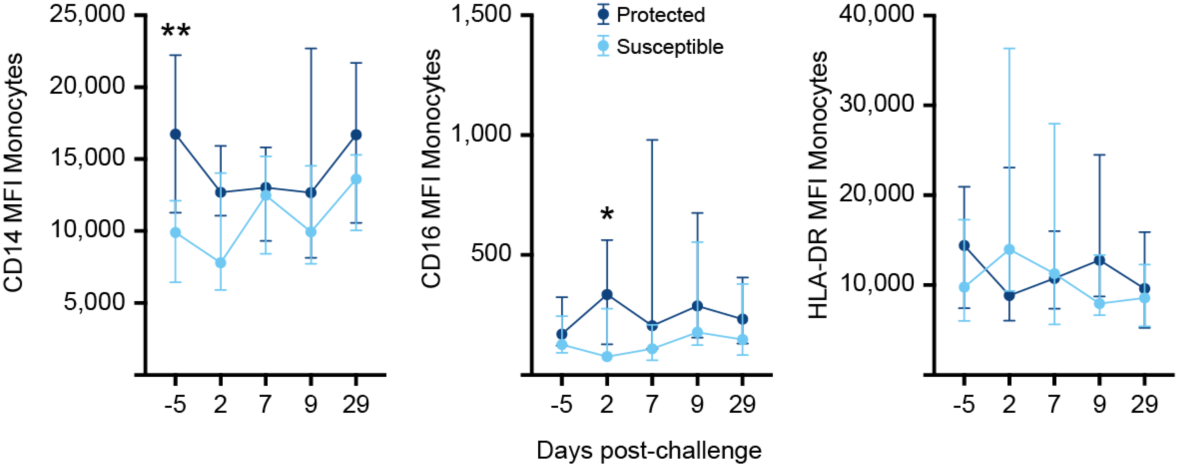
Monocytes are not recruited into the nasal mucosa after pneumococcal challenge. **a**, Line graph (median and interquartile range) of expression levels of CD14, CD16, and HLA-DR on monocytes before (day –5) and after challenge (day 2, day 7, day 9 and day 29) in protected (dark blue, n=25) and susceptible (light blue, n=14) older adults. The expression level of CD14 was significantly lower on nasal monocytes at baseline (Mann Whitney U test, baseline: p=0.0019).

## Discussion

Exposure to respiratory pathogens occurs in the upper respiratory tract and immunological responses here are critical for either the establishment or clearance of infection. Failure to clear infection increases the risk of microaspiration of pathogens into the lung and development of pneumonia. Older adults are at an increased risk of pneumococcal disease although rates, duration and densities of *S. pneumoniae* colonization in the upper respiratory tract are comparable or lower than those observed in younger adults^16,29^. Age has been associated with cellular senescence which in turn drives “inflammaging” akin to chronic inflammation with the release of proinflammatory cytokines and increased vascular permeability^14^. Aging alters the innate immune response with changes in neutrophil recruitment into tissue, reduces phagocytosis and NETosis and reduced phagocytosis and antigen presentation by macrophages and dendritic cells (reviewed in ^30^). An alternative and not necessarily contradictory view is that changes in cellular composition and response to recall antigens during aging are appropriate adaption to life-long exposure to pathogens with “inflammaging” as an early sign of underlying disease independent of age ^31^. Here we investigated the immune response in nasal tissue before and after pneumococcal controlled human infection in healthy older adults and compared results with existing data of younger adults^9^. Remarkably, we observed that older adults who became colonized presented with a susceptibility profile both at the transcriptional and cellular level before pneumococcal challenge.

Analysis of the nasal transcriptome revealed overexpression of genes associated with the reactome pathway “neutrophil degranulation” in older susceptible adults compared to those protected from pneumococcal colonization. Granulocytes, the majority of which are neutrophils, are abundant in the nasal mucosa and the first line of defence against bacterial infection through mechanisms such as antibody-mediated phagocytosis, production of radical oxygen species, degranulation and NETosis. While the frequency of granulocytes remained unchanged before and after pneumococcal challenge in older study participants, expression levels of genes associated with the reactome pathway “neutrophil degranulation” positively corelated with MPO concentration in nasal lining fluid and negatively correlated with CD16 expression levels on granulocytes, both of which indicate higher activation of neutrophils in the nasal mucosa of susceptible adults before pneumococcal challenge. Although older adults had higher levels of MPO compared to younger adults before pneumococcal challenge, there were no differences between susceptible and protected older adults and were maintained at comparable level after challenge. It has been reported that with age, the microbiome of the nasal and oral cavity is no longer distinct^32^ but whether this contributes to increases risk of disease is unknown. Although the exact driver of increased activation of neutrophils in susceptible older adults we report here require further investigation, it may result in local tissue damage which in turn may increase adhesion and micro-invasion of pneumococcus in the nasal mucosa^33^.

It is also noteworthy that the *FCAR* gene encoding the FcαR was 3-fold overexpressed in susceptible adults before pneumococcal challenge. Although anti-pneumococcal secretory IgA is cleaved by the *S. pneumoniae* IgA1 protease (ZmpA), it is an important opsonin for other viral and bacterial pathogens. Gene expression levels of *FCRG3A* and *FCGR3B*, encoding FcγRIIIA expressed on NK cells and monocytes and FcγRIIIB expressed on neutrophils, increased two days after pneumococcal challenge in susceptible study participants, although increased expression of CD16 (FcγRIII) was not evident at the cellular level. It has been reported in other studies^34,35^ that CD16 expression is reduced on neutrophils from older adults and associated with reduced phagocytosis of pneumococcus. We observed that CD16 expression was generally lower in susceptible study participants only both before and after pneumococcal challenge. CD16 is a GPI-anchored protein and rapidly shed from the surface of granulocytes upon activation or apoptosis which may explain the discrepancy between transcriptional and cellular data. CD16-mediated signalling upon crosslinking with immune complexes depends on association of CD16 with CD32 (FcγRII). Shedding of CD16 may result in better accessibility of FcγRIIA (CD32) on the surface of neutrophils enhancing subsequent Fc-mediated phagocytosis or extracellular killing of bacteria^36^. Although the role of FcR-mediated phagocytosis by neutrophils in the nasal mucosa has to be confirmed at the functional level, we speculate that increased expression of Fc-receptors on nasal neutrophils of susceptible older adults after colonization onset may contribute to the clearance of pneumococci.

In our study, the concentration of both CXCL10 and the related cytokine CXCL9 were increased at the protein level and transcriptional level in older susceptible adults compared to protected older adults before pneumococcal challenge. CXCL10 and the closely related CXCL9 are largely induced by IFNγ and chemoattractants for NK– and T cells, important for the clearance of viral infections. Increased concentrations of CXCL10 have been reported in nasal lining fluid before challenge with RSV in those who developed cold symptoms^37^ and younger study participants receiving the nasal live attenuated influenza vaccine (LAIV) or with asymptomatic viral infection before pneumococcal challenge were associated with increased pneumococcal load^9^. In a recent study evaluating the use of high levels of CXCL10 in nasal lining fluids as a marker for undiagnosed viral infections, the authors detected a distinct profile of high levels of CXCL10 together with neutrophilic inflammation in bacterial and viral co-infections^38^. However, it is unlikely that the increased CXCL10 concentration in our study was due to viral infection because individuals with an asymptomatic viral infection were excluded from our analysis. The role of CXCL10 in bacterial infections is less clear. Increased CXCL10 concentrations were also associated with pneumonia in animal models of pneumococcal disease and it is also a promising marker which distinguished between CAP caused by *Staphylococcus aureus* or *S. pneumoniae* with higher levels detected in pneumococcal CAP^39^. Furthermore, CXCL10 can be produced by tissue-infiltrating neutrophils which express its receptor CXCR3 and acts on these neutrophils in an autocrine fashion followed by degranulation, ROS production and chemotaxis thus contributing to airway inflammation^40^.

High serum concentrations of CXCL9 are a strong predictor of cellular aging and cardio-vascular health even in healthy adults^41^. It is produced by endothelial cells, macrophages and neutrophils^42^. Whether CXCL9 is indicative of neutrophilic inflammation or produced by endothelial cells in the nasal mucosa in our study remains unclear but given its role in biological aging, its association with increased susceptibility to colonization with *S. pneumoniae* requires further investigation and if confirmed may offer an opportunity to identify older adults at greater risk of pneumococcal infection.

Another important observation was the lack of recruitment of monocytes into the nasal mucosa after pneumococcal challenge unlike younger adults^9^. It is not known whether CCL2 is not induced or expression of the CCL2 receptors CCR2 and CCR4 is impaired. However, both CCL2 and CCR2 expression on monocytes are crucial for additional recruitment of and clearance of pneumococcus by monocytes^43^. A recent study in a mouse model pneumococcal pneumonia suggested that age is associated with higher expression of CCR2 on monocytes but reduced phagocytosis of *S. pneumoniae*^44^. Therefore, it will be important to analyse the phenotype and function of monocytes over the course of colonization in an experimental infection model in more detail, together with the frequency and function of dendritic cells, both in healthy younger and older adults since both populations play a role in the activation of adaptive immune responses both in the nasal mucosa and systemically.

Given that in our study population, older adults cleared pneumococcal infection from the nasal mucosa without showing higher bacterial densities or any discernible delay in colonization duration, the question arises how the above changes in the nasal mucosa at the cellular and transcriptional level reflect the increased risk of older adults to develop pneumococcal disease.

Although the upper and lower respiratory tract differ in composition in both type and frequency of specialised epithelial cells and immune cells, there is sufficient overlap between compartments as assessed at the single cell level scRNAseq^45–47^. One possible explanation is that older adults have physiological changes such as reduced mucocillary clearance and increased expression of host receptors binding *S. pneumoniae* that leads to increased micro-aspiration into their lungs^48–51^. Micro-aspiration of pneumococcus into the lung may result in more pronounced inflammation with increased tissue damage if neutrophils are recruited or activated in a similar manner to those observed in the nasopharynx. Neutrophils in the lung are found in increased frequency in the lung and contribute to inflammation during bacterial and viral infections^52–54^. In addition, adults at risk of pneumonia have a delay in lung efferocytosis – the phagocytic clearance of apoptotic cells – following community acquired pneumonia^55^. In a mouse model of pneumococcal and viral co-infection, older mice showed rapid activation of neutrophils in the lung in the absence of increased bacterial burden suggesting that dysregulation of immune responses are a major factor contributing to pneumonia^56^. Understanding the role of individual pathogenic bacteria such as pneumococcus and its relationship with activation (or lack of) of immunological pathways that could result in increased susceptibility to pneumococcal disease are a prerequisite for future studies analysing in detail the interaction between bacterial and viral infection in clinically vulnerable populations, and thus improving diagnosis and treatment.

## Online Methods

### Study Design & Demographics

Healthy adults aged 50-84 were inoculated with an estimated 80,000 CFU per nostril of *Streptococcus pneumoniae* serotype 6B (strain BHN418 (29), GenBank accession number ASHP00000000.1) as previously described^7,57^. Key eligibility criteria included capacity to give informed consent, no immunocompromised state or contact with susceptible individuals or children, no pneumococcal or influenza vaccine or infection in the last 2 years and not having taken part in EHPC studies in the past 3 years. All volunteers gave written informed consent, and research was conducted in compliance with all relevant ethical regulations. Participants who carried non-experimental pneumococcal strains at baseline (day –5) or had a viral infection were excluded from all analyses.

Ethical approval was provided by the East Liverpool National Health Service Research and Ethics Committee (reference numbers 15/NW/0146 and 14/NW/1460) and Human Tissue Authority licensing number 12548.

Detailed demographic data of older adults >50yrs can be found in Adler 2021^16^. For comparison, data from younger adults who were part of a control group of an Influenza co-infection study were re-analysed^9,18^. Some data on baseline (day –5) nasal cell populations before inoculation with Spn6B were presented in a pre-print^58^ but the analysis of baseline nasal cell data presented here is sufficiently different and substantially extends the data presented in the preprint.

### Determination of carriage

Nasal washes were performed at day –5 and on days 2, 7, 9, 14, 22 and 29 post-challenge and colonization determined by the presence of pneumococcus in nasal wash samples at any time point post-challenge up to and including day 29, detected using classical microbiology^7,57^.

### Nasal curettage

Nasal cells were collected on days –5, 2, 7, 9 and 29 by scraping the inferior turbinate from participants consecutively using four curettes (Rhino-pro®, Arlington Scientific), as described previously^25^. Two curette samples were collected into PBS + 5mM EDTA and 0.5% heat inactivated FBS for immunophenotyping and an additional two curette samples were collected into RLT (Qiagen) and stored at –80°C for transcriptome analysis.

### Nasal Cell Immunophenotyping

Cells were dislodged from curettes and processed immediately. Briefly, the nasal cells were stained for 15 minutes with LIVE/DEAD® Fixable Violet Dead Cell Stain (Thermofisher) before the addition of a cocktail of monoclonal antibodies for 20 minutes at 4°C protected from light (Supplementary Table S7) Following incubation, cells were washed (440g for 5 min) with 3mL PBS, filtered through a 70µm mesh filter (ThermoFisher) and resuspended in 200µL Cell FIX™ (BD Biosciences). Samples were acquired on a LSR II cytometer (BD Biosciences) and analysed using Flowjo software version 10 (Treestar). Compensation matrices were set using compensation beads (BD Biosciences and Thermofisher). Samples with less than 500 CD45+ leukocytes or 250 EpCam+ epithelial cells were excluded from analysis. To adjust for the variability of cells collected within and between study participants, absolute cell counts of the target populations were normalized with the absolute number of epithelial cells. The gating strategy is shown in Supplementary Fig. 5.

### Anti-6B capsular polysaccharide IgG

Anti-6B CPS IgG titres were measured using a modification of the WHO enzyme-linked immunosorbent assay (ELISA) protocol. Nasal wash (NW) samples and reference serum 89SF (US Food and Drug Administration (FDA)) were depleted of cell wall polysaccharide (CWPS) antibodies by pre-absorption with 10µg/mL solution of CWPS (Statens Serum Institut, Copenhagen, Denmark) in phosphate-buffered saline (PBS) blocked with heat-inactivated foetal bovine serum (ThermoFisher, Basingstoke UK) for 30 minutes. 96-well plates (Maxisorp microtiter, Nunc, Roskilde, Denmark) were coated with 5µg/mL purified pneumococcal 6B capsular polysaccharide (Statens Serum Institut) overnight at 4°C, washed and blocked with 1% BSA. Reference serum 89SF was serially diluted as a standard curve. Samples and standards were transferred to plates and incubated for two hours at room temperature, followed by three washes with PBS/0.05% Tween (PBS-T), before incubation with the secondary antibody diluted 1:4000 (A9544; Sigma-Aldrich Corporation, Dorset, UK) for 90 minutes. Wells were washed as before and developed by incubation with p-nitrophenylphosphate (Sigma-Aldrich Corporation) for 15-20 minutes at room temperature. Optical densities were measured at 405nm using a FLUOstar Omega plate reader (BMG Labtech GmbH, Ortenberg, Germany). Samples were analyzed in duplicate, and samples with a coefficient of variation (CV) of > 25% were excluded.

### Nasal Fluid Multiplex Cytokine Measurement

Nasal lining fluid was collected using nasosorption filters (Nasosorption, Hunt Developments) and stored at –80°C. For detection of cytokines in the nasal lining fluid, nasosorption samples were eluted in 100μl assay buffer (Thermo Fisher Scientific) by centrifugation at 2800g for 10 minutes. The eluate was cleared by further centrifugation at 16,000g for 10 min. Samples were analysed on an LX200 using a 30-plex magnetic human Luminex cytokine kit (LHC6003M; Thermo Fisher Scientific) and analysed with xPonent3.1 software following the manufacturer’s instructions. Samples were analysed in duplicate, and samples with a coefficient of variation (CV) of > 25% were excluded.

### Myeloperoxidase ELISA

Levels of myeloperoxidase in the nasal wash were determined using the Human Myeloperoxidase DuoSet ELISA Kit (R&D Systems, Abingdon, UK) following manufacturer’s instructions. Briefly, 96-well plates (Maxisorp microtiter, Nunc, Roskilde, Denmark) were coated with 4μg/mL capture antibody overnight and blocked with 1% BSA in PBS for 1 hour at room temperature. Samples and standards were incubated in assay plates for 2 hours at room temperature. Detection antibody (50ng/ml) was added to the wells for 2 hours at room temperature, followed by 20 minutes incubation with Streptavidin-HRP (1:200; Fisher Scientific) at room temperature. The plates were developed using TMB-Turbo Substrate (Fisher Scientific) for 20 minutes and the reaction stopped by adding 2N H_2_SO4 in a 1:1 ratio. Optical density was measured at 450nm and corrected for optical imperfection (540nm). All samples were run in duplicate with a CV <25%.

### Nasal Cell RNA Sequencing and Bioinformatic Analysis

*RNA sequencing*: Nasal cells were collected in RLT (QIAGEN) and stored at –80°C until extraction. RNA extraction (RNeasy; QIAGEN), sample integrity assessment (Bioanalyzer; Agilent 2100), library preparation and RNA sequencing (BGISEQ-500RS) were performed at the Beijing Genome Institute. Quality control of raw sequencing data was done using FastQC tool. Mapping to a human reference genome assembly (GRCh38) was done using STAR 2.5.0a^59^. Read counts from the resulting binary alignment map (BAM) files were obtained with featureCounts^60^ using a general transfer format (GTF) gene annotation from the Ensembl database^61^The R/Bioconductor package DESeq2 (version 1.34.0) was used to identify differentially expressed genes among the samples after removing absent features (zero counts in more than 75% of samples)^62^. Genes with a p-value of < 0.05 and an absolute fold-change of > 2.0 were identified as differentially expressed. The complete set of raw sequences were deposited at the NCBI through BioProject (PRJNAXX).

*Functional analysis*: We performed two functional analysis approaches regarding the biological pathways related to ageing or carriage status. Gene Set Variation Analysis (GSVA) was used to identify pathways independent of comparison between groups of samples using R package gsva (version 1.42.0) and log_2_ count per million (CPM) matrix count, and the parameters kcdf=”Gaussian”, min.sz=15, and max.sz=500. Gene Set Enrichment Analysis (GSEA) was used to identify which pathways were represented in the ranked transcriptome (log_2_ fold-change), independently of the DE cut-off^63^ using R package fgsea (version 1.20.0). The criteria used for statistical enrichment for GSEA were a p value < 0.05 and enrichment of at least five genes for each pathway. The database used for functional analysis was Reactome, release 79 and Pathway Browser version 3.7^50^.

*Co-expression analysis*: For co-expression analysis, counts were normalized using log_2_ counts per million (CPM), and the log_2_ fold-change was calculated for each time point in a subject-wise manner. The co-expression analysis was performed separately for each group (older and younger, using baseline, day 2, and day 9 after pneumococcal challenge and susceptible and protected samples) using the R /Bioconductor package CEMiTool (version 1.18.1)^19^. This package unifies the discovery and the analysis of co-expression gene modules, evaluating whether modules contain genes that are over-represented by specific pathways or altered in a specific sample group. A p-value = 0.05 was applied for filtering genes with low expression levels.

*M2 CEMiTool module protein-protein interaction network*: To identify the relationship between genes (protein-protein interaction) from M2 CEMiTool modules, we filtered which genes were differentially expressed at least one comparison (susceptible vs protected at baseline, day 2 or day 9 after challenge), and we used the networkanalyst.ca web-based tool (version 3.0). We used the “Gene List Input” option, a list of M2 DE gene symbols and StringDB^64^ to find the genes interaction evidence under parameters, confidence score cutoff = 900, not required experimental evidence and zero-order network to use just genes from the input list.

*Deconvolution analysis*: Cell type estimation was performed using the CIBERSORTx web-based tool^65^, with log_2_ count per million (CPM) gene expression and single-cell atlas data from sorted monocytes cells^66^ and bone marrow and mature neutrophils bulk sequencing^67^ from healthy human donors. Both studies were used as a reference to predict cell type proportion.

### Statistical Analysis

To identify the correlation between a pair of features, we first imputed the missing data using the R package missForest (version 1.5) and then calculated the Spearman rho coefficient and p values using the R base function cor.test.

Two-tailed statistical tests were used throughout the study. For comparison between study groups, Mann Whitney test was used. For comparison of study participants within a group over time, Friedmann test with Dunn Hochberg correction was used. P values were considered significant at <0.05 unless stated otherwise. Data were analysed using GraphPad prism (version 9) or R studio (2021.09.2 Build 382) and R (version 4.1.2).

## Data Availability

Raw RNA sequencing data have been deposited in the Gene Expression Omnibus repository. All other underlying data are provided in the manuscript.

## Acknowledgements

We thank all study participants for their contribution and generous donation of time and tissue samples. We acknowledge the invaluable contribution of the clinical and laboratory team collecting and processing samples. This study was funded by the Melinda Gates Foundation (grant OPP1117728); the UK Medical Research Council (grant MR/M011569/1).

---

**Fig. S1:**
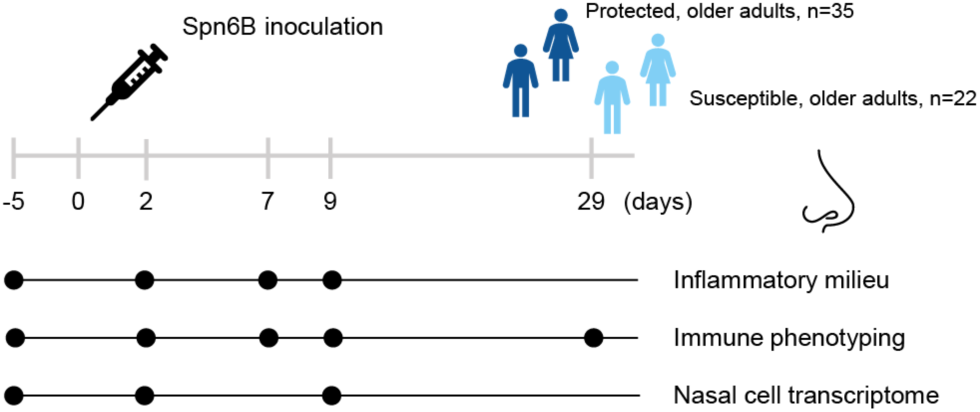
Study design. Study design showing inoculation of study participants with Spn6B at day 0 and sample collection before (day –5) and after inoculation with Spn6B (day 2, day 7, day 9, and day 29). Samples collected were nasal lining fluid (nasosorption samples) for determination of the nasal inflammatory milieu and nasal cells (nasal curettage) for immune phenotyping and analysis of the nasal transcriptome. Study participants who remained protected (n=35) are depicted in dark

**Fig. S2.**
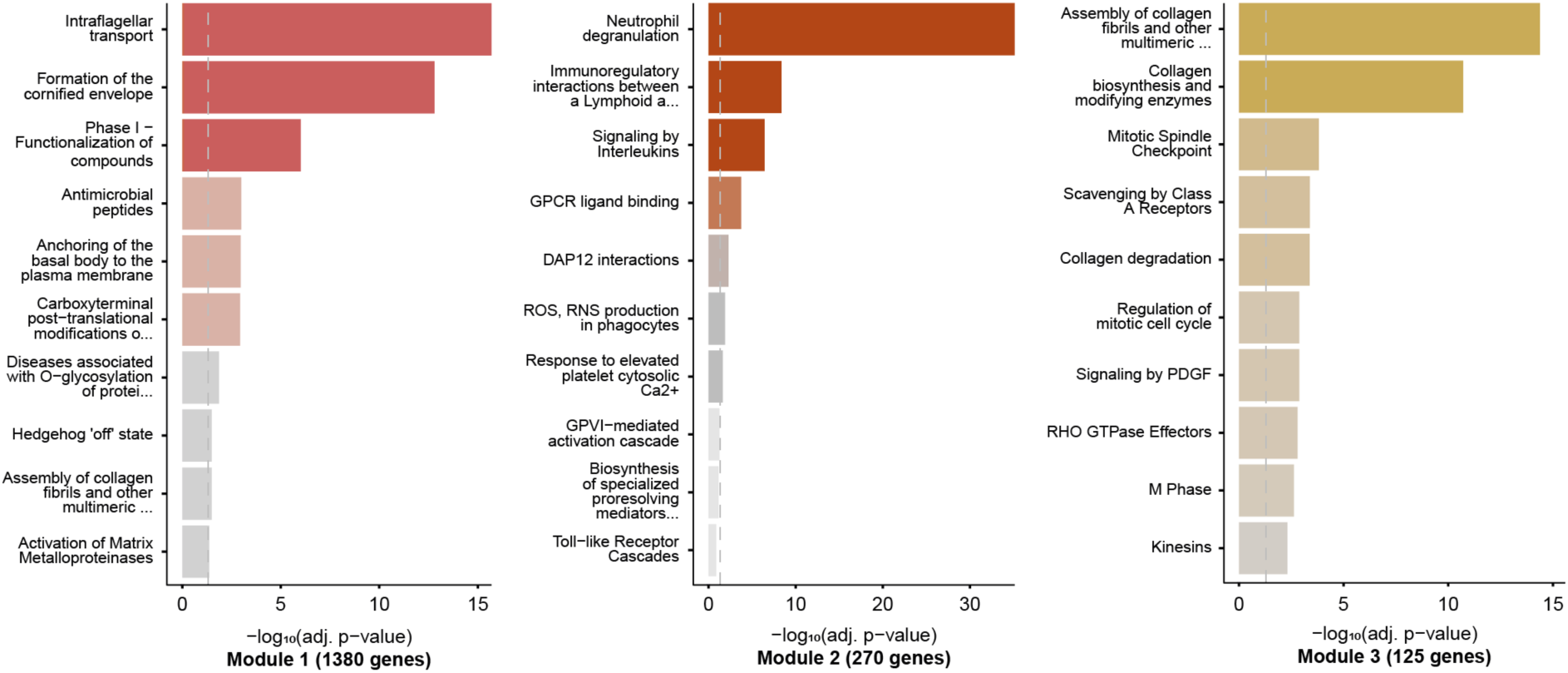
Modules of co-expressed gene sets functional analysis. Over-Representation Analysis (ORA) was used to determine biological pathways (using Reactome level 3 gene sets) of co-expressed genes across all modules identified by CEMiTool.

**Fig. S3.**
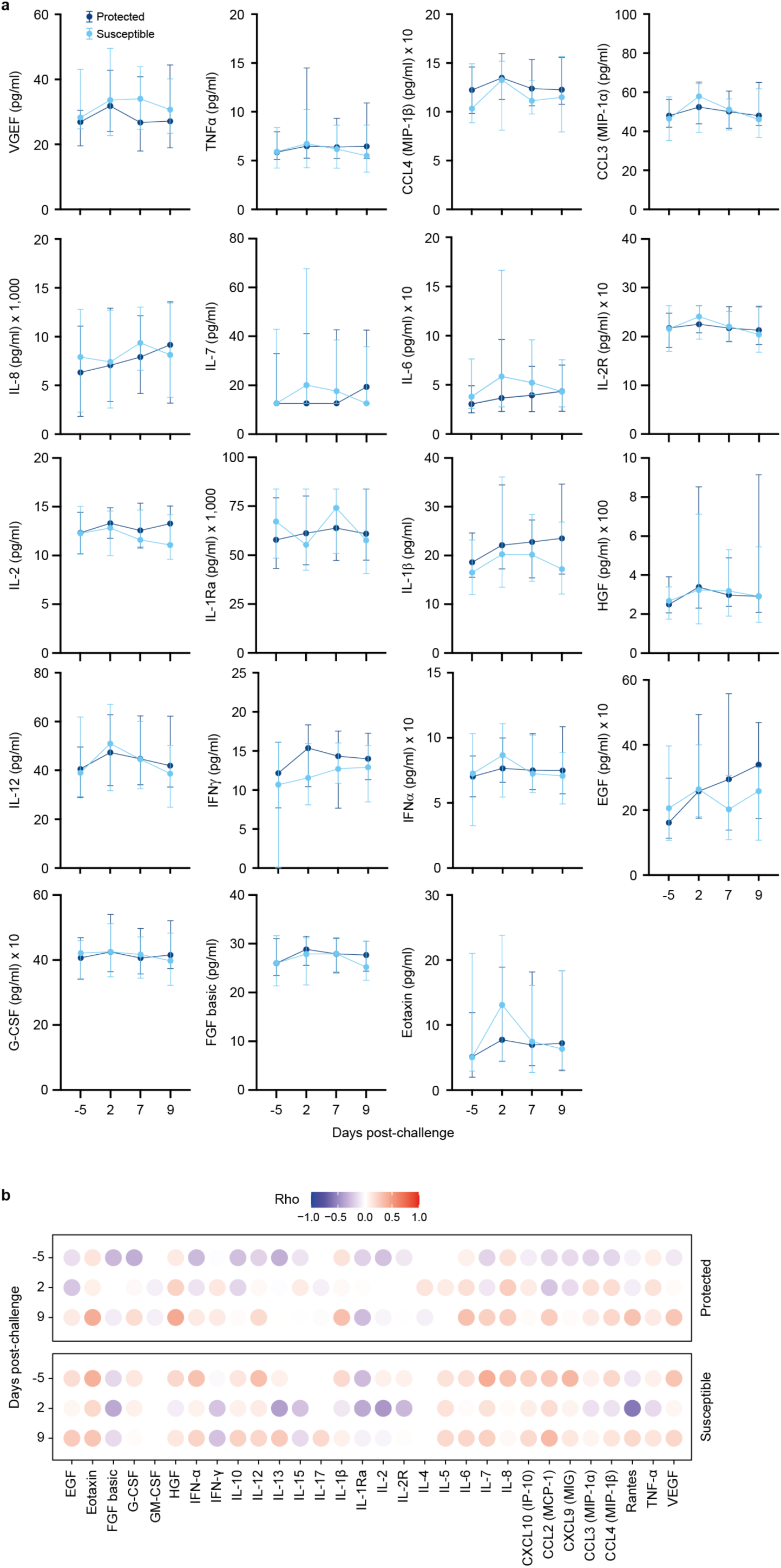
Cytokine concentration in the nasal lining fluid of older adults experimentally challenged with *S. pneumoniae* 6B. **a**, Line graphs showing the median and interquartile range of cytokine concentrations in the nasal lining fluid in older study participants before (day –5) and after (day 2, day 7 and day 9) pneumococcal challenge in susceptible (light blue line, n=22) and protected (dark blue line, n=35) study participants. **b**, Correlation plot of association between age of protected and susceptible study participants with cytokine concentration before (day –5) and after (day2 and day9) pneumococcal challenge. Correlation was tested using Spearman rho with Benjamin Hochberg adjustment for multiple comparison.

**Fig. S4.**
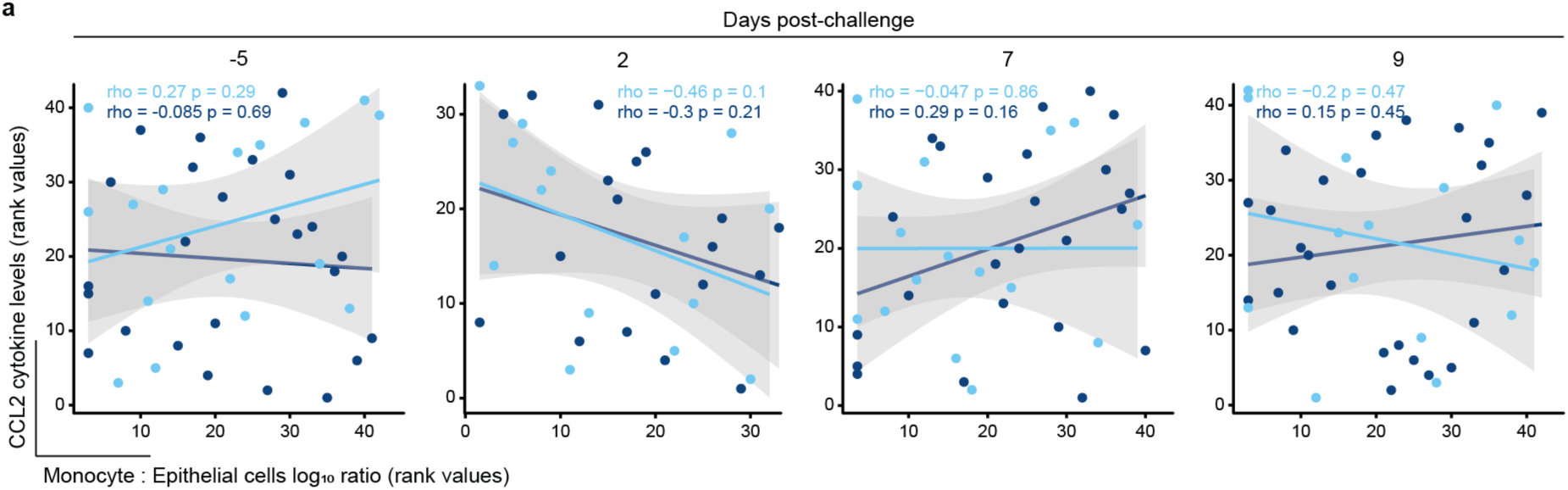
Monocytes are not correlated with CCL2 levels into the nasal mucosa before and after pneumococcal challenge. Scatter plot of the CCL2 level (pg/ml) and monocyte: epithelial cells ratio before (day –5) and after challenge (day 2, day 7, and day 9), which showed no correlation in susceptible (light blue dots) and protected (dark blue dots) study participants.

**Fig. S5.**
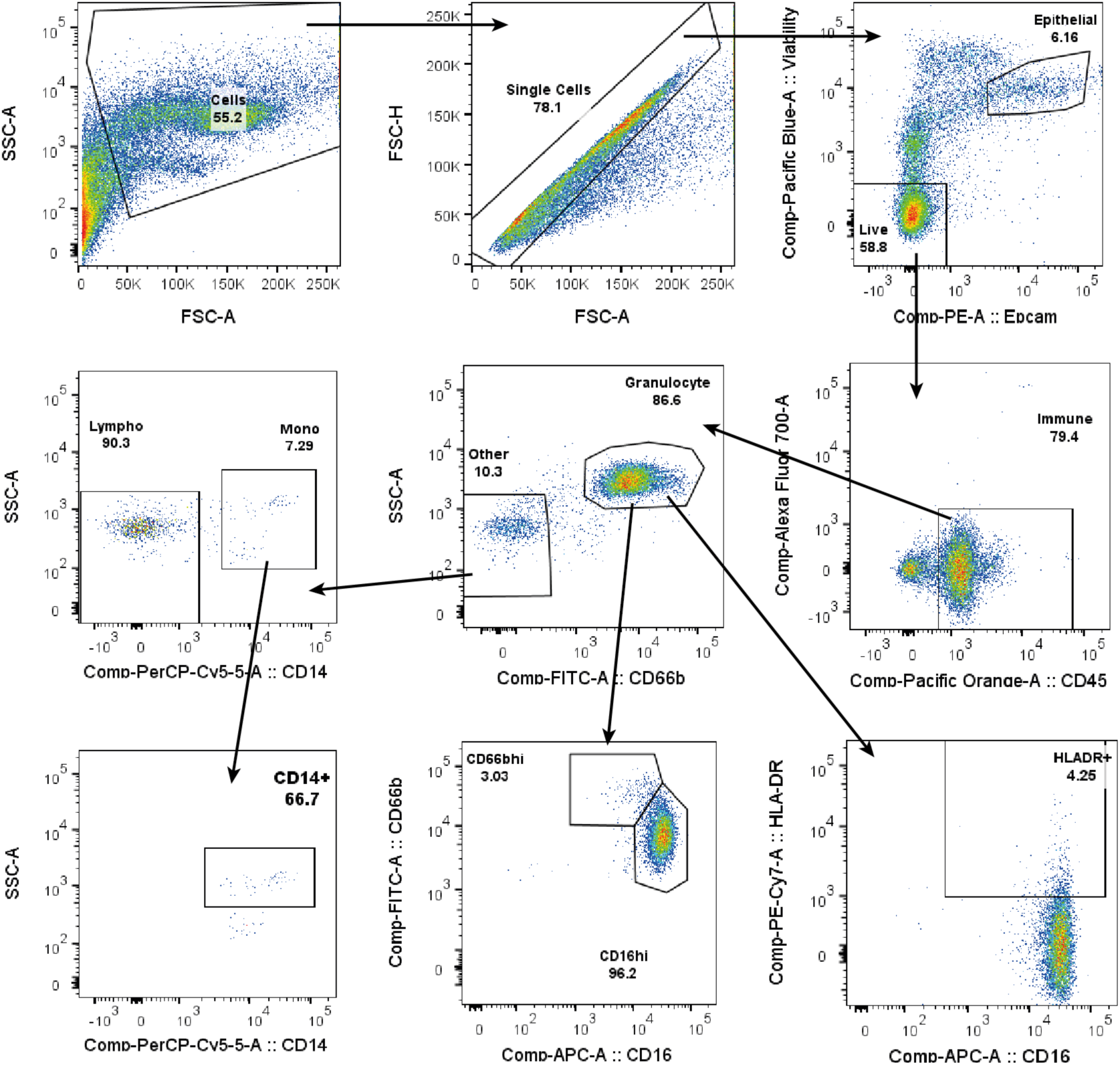
Gating strategy for nasal cells of one representative study participant.

